# Machine Learning for Integrating Social Determinants in Cardiovascular Disease Prediction Models: A Systematic Review

**DOI:** 10.1101/2020.09.11.20192989

**Authors:** Yuan Zhao, Erica P. Wood, Nicholas Mirin, Rajesh Vedanthan, Stephanie H. Cook, Rumi Chunara

## Abstract

**Background:** Cardiovascular disease (CVD) is the number one cause of death worldwide, and CVD burden is increasing in low-resource settings and for lower socioeconomic groups worldwide. Machine learning (ML) algorithms are rapidly being developed and incorporated into clinical practice for CVD prediction and treatment decisions. Significant opportunities for reducing death and disability from cardiovascular disease worldwide lie with addressing the social determinants of cardiovascular outcomes. We sought to review how social determinants of health (SDoH) and variables along their causal pathway are being included in ML algorithms in order to develop best practices for development of future machine learning algorithms that include social determinants.

**Methods:** We conducted a systematic review using five databases (PubMed, Embase, Web of Science, IEEE Xplore and ACM Digital Library). We identified English language articles published from inception to April 10, 2020, which reported on the use of machine learning for cardiovascular disease prediction, that incorporated SDoH and related variables. We included studies that used data from any source or study type. Studies were excluded if they did not include the use of any machine learning algorithm, were developed for non-humans, the outcomes were bio-markers, mediators, surgery or medication of CVD, rehabilitation or mental health outcomes after CVD or cost-effective analysis of CVD, the manuscript was non-English, or was a review or meta-analysis. We also excluded articles presented at conferences as abstracts and the full texts were not obtainable. The study was registered with PROSPERO (CRD42020175466).

**Findings:** Of 2870 articles identified, 96 were eligible for inclusion. Most studies that compared ML and regression showed increased performance of ML, and most studies that compared performance with or without SDoH/related variables showed increased performance with them. The most frequently included SDoH variables were race/ethnicity, income, education and marital status. Studies were largely from North America, Europe and China, limiting the diversity of included populations and variance in social determinants.

**Interpretation:** Findings show that machine learning models, as well as SDoH and related variables, improve CVD prediction model performance. The limited variety of sources and data in studies emphasize that there is opportunity to include more SDoH variables, especially environmental ones, that are known CVD risk factors in machine learning CVD prediction models. Given their flexibility, ML may provide opportunity to incorporate and model the complex nature of social determinants. Such data should be recorded in electronic databases to enable their use.

**Funding:** We acknowledge funding from Blue Cross Blue Shield of Louisiana. The funder had no role in the decision to publish.

## Introduction

An estimated 17.9 million people die each year from cardiovascular diseases (CVD), which represent 31% of all deaths worldwide and the number one cause of death.^1^ Low-income and middle-income countries carry 75% of the burden of CVD deaths worldwide and in high-income countries, lower socioeconomic groups have a higher incidence of CVD and higher mortality due to CVD.^1,2^ In high-income countries such as the United States, the prevalence of CVD is expected to rise 10% between 2010 and 2030,^3^ not only in the aging population but also notably via stark disparities among socioeconomic and racial groups.^4,5^ Direct causes for these shifts in CVD burden have been well-studied, attributed to changes in diet (increased consumption of processed foods)^6^ and physical activity (more sedentary lifestyles),^7^ resulting in a dramatic rise in conditions such as obesity, hypertension, and diabetes mellitus. These changes are shaped by the “conditions in which people are born, grow, live, work and age”, referred to by the World Health Organization as social determinants of health (SDoH).^8^

Multinational, prospective cohort studies as well as ecologic analyses have shown that SDoH contribute to over 35% of the population attributable risk of various cardiovascular diseases,^9,10^ among which education, income and occupation are particularly influential.^11^ Research has also illuminated mechanisms of action; social factors usually interact with each other through the mediation of or effect modification by psychological and biological pathways, exerting a long-term effect on cardiovascular outcomes.^5,12^ Social determinants also result in unequal sharing of the benefit of advances in CVD prevention and treatment.^13^ Given the critical importance of social determinants with respect to disease risk, it is clear that better capturing the interaction and relative influence of such factors in relation to traditional CVD risk factors of hypertension, diabetes and hyperlipidemia provides the most significant opportunity to reduce CVD burden.^5,11,12,14^

Meanwhile, artificial intelligence (AI) and machine learning (an application of AI for detecting patterns from data)^15^ tools have started to be adopted in clinical research, prompted by recent progress in advanced computing strategies as well as the proliferation of electronic medical record databases.^16^ Machine learning methods have demonstrated improvement across multiple metrics for prediction of CVD risk, incidence and outcomes^17-19^ over traditional risk scores such as those from the American College of Cardiology or American Heart Association.^20^ As a data-driven approach, machine learning provides more flexibility in modeling complex relationships between predictors, which can be particularly advantageous in addressing the multi-level interactions between different social determinants and CVD outcomes, as well as uncovering novel risk factors. Though the increased flexibility of machine learning models is appealing, given the rapid rise of machine learning approaches including studies which incorporate social determinants, we need to better understand best practices for such modelling approaches for CVD risk prediction particularly in the context of those including SDoH.

Thus, we performed a systematic review to understand the current landscape of how social determinants are being used in machine learning models for CVD prediction. Specifically, we sought to examine which types of machine learning algorithms and types of social determinant variables are being used, and for which populations. Indeed, understanding the manner in which SDoH are incorporated into such models is critical in order to tease apart the distinct the biological and social influences, along with their interactions, that make populations different and in need of a different standard of care. Findings from this review serve to inform the design of future machine learning approaches and identify areas for methodological innovation in order to improve early prediction of CVD and reduce its significant disease burden.^21,22^

## Method

### Search strategy and selection criteria

First, YZ with the help of an expert librarian, did a comprehensive search of five databases: PubMed, Embase, Web of Science, IEEE Xplore and ACM Digital Library on April 10th, 2020, to identify all relevant articles on machine learning integrating social determinants in cardiovascular disease prediction models published in English. IEEE Xplore and ACM Digital Library were included specifically to comprehensively capture computer science articles related to our review. Papers from inception until the search date were included. To ensure the quality of included papers, we only included peer-reviewed articles published on journals or accepted in conferences and excluded non-peer reviewed grey literature or arXiv/medRxiv papers.

We identified the terms of social determinants of health (SDoH) using the broader definition from the World Health Organization and Center for Disease Control and Prevention Healthy People 2020 initiative which delineates SDoH in five key areas: economic stability, education, social and community context (e.g. “race/ethnicity”, “income” and “education”), health and health care, neighborhood and built environment (e.g. “living environment”, “pollution” and “residence characteristics”).^23^ Figure 1 is a socio-ecological conceptual model adapted from Healthy People 2020, the United States federal government’s national health agenda,^24^ which illustrates the multifactorial nature of social-ecological influences on health. The framework emphasizes the existence of proximate, or “downstream,” health influences (e.g., smoking) that are shaped by distal, or “upstream,” factors (e.g., social norms regarding smoking, tobacco regulations). Therefore, for a robust review, we also included prominent factors including health-related behaviors along the causal pathway (e.g. “diet”, “smoking” and “physical activity”), as although these are enacted at the individual level, they are shaped at social and economic levels.^25^ This enables us to understand comprehensively how social determinants and factors they directly shape are assessed in relation to CVD. Age, gender and race are also in the causal pathway.^26^ BMI was also included as it is influenced by social factors and causes diabetes which directly affects CVD.^27^

**Figure 1:**
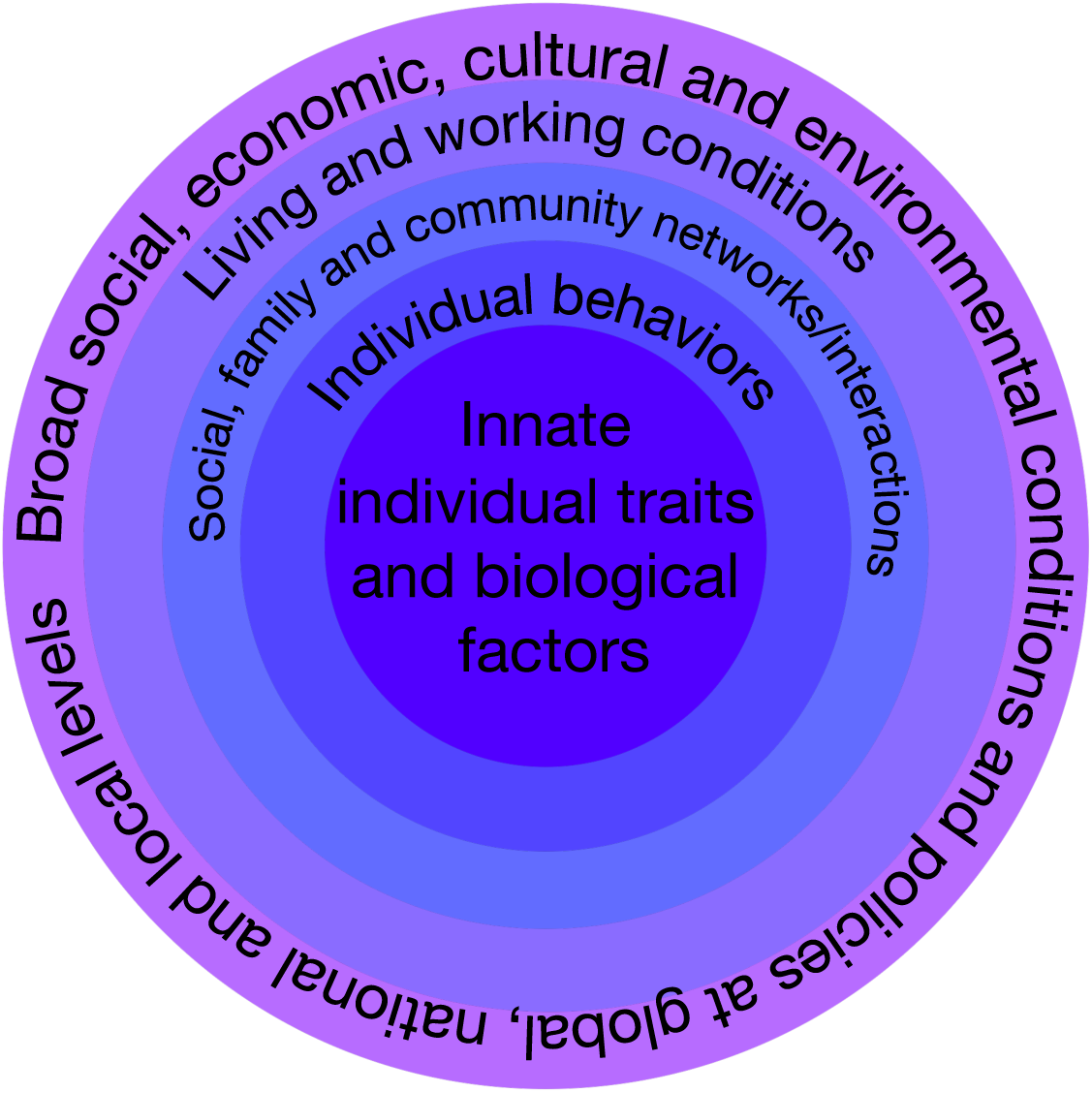
Socio-ecological framework of health; conceptual model used in the study, adapted from Healthy People 2020, the United States federal government’s national health agenda

For search terms related to machine learning, we included all commonly used supervised machine learning methods. Supervised machine learning algorithms are those that perform reasoning (i.e. prediction) from observations of the features (e.g. clinical data, social determinants) based on externally supplied examples which include the features linked to outcome “labels” (e.g. CVD outcomes). Thus supervised machine learning was a focus as the types of tasks considered in the literature usually utilized labeled outcomes of CVD.^28^ Commonly used unsupervised machine learning algorithms captured by the search were also included in the abstract and full text screening to ensure all types of possible studies were considered. We also added search terms to capture deep learning and ensemble methods as they are widely used in current clinical research.^29^

The search terms for CVD outcomes included cardiovascular ischemic outcomes, coronary heart disease and cerebrovascular disease which are caused by atherosclerotic cardiovascular disease (ASCVD). These cardiovascular diseases cause the highest mortality, and estimated years of lives lost attributed to these conditions have increased in recent years.^12,30^ For each of the key areas of social determinants and included variables, machine learning and CVD, we identified keywords by referencing previous review papers on social determinants and cardiovascular diseases,^11,31^ related studies of different social determinants^31-35^ or consulting experts to include relevant concepts. Full search strategies are provided in the appendix.

Once papers were identified via the search terms, all study designs and all populations were included if the article utilized any SDoH or health behaviors as features in the machine learning models (in addition to age and gender, as we found that these were commonly included as standard practice and not specifically to represent their contribution as social determinants) were deemed eligible. Eligibility was also considered if the outcomes were CVD-related, including incidence, survival, mortality, hospital admission and readmission etc. We did not restrict time of publication to enable capturing the trend of these types of papers over time. Studies were excluded if they did not include the use of any machine learning algorithm, were developed for non-humans, the outcomes were bio-markers, mediators, surgery or medication for CVD, rehabilitation or mental health outcomes after CVD diagnosis or cost-effectiveness analysis of CVD treatment, the manuscript was non-English, or was a review or meta-analysis. We also excluded articles presented at conferences as abstracts and the full texts were not obtainable. This review was registered with PROSPERO (CRD42020175466) and conducted in accordance with the Preferred Reporting Items for Systematic Reviews and Meta-Analyses (PRISMA) method. To supplement the bibliographic database searches, we also used Google Scholar to scrutinize all keywords regarding their relevance in articles as well as examine potential articles to identify if they were eligible. Duplicates were removed in the process.

Three investigators (YZ, EPW, and NM) screened the title and abstract: each article retrieved was independently assessed by two reviewers to determine its eligibility for full-text review. Conflicts were resolved by discussion and validation from a third reviewer. After initial appraisal, we retrieved full texts of eligible articles.

### Data analysis

Data were extracted from individual articles independently by two reviewers (of YZ, EPW, and NM) and checked by the third reviewer according to criteria in a standardized extraction form. All data extraction was cross-checked, and disagreements were resolved by discussion or referral to the third reviewer. Information extracted included year of publication, country, population, social determinants included in the machine learning algorithm, machine learning algorithms, cardiovascular disease outcomes, data source and performance of the algorithms. For each article, we defined several criteria to assess the quality of the study based on best practices in machine learning^36^ including (1) whether machine learning model performance was evaluated; (2) whether a hyperparameter (a parameter whose value is used to control the learning process) tuning process was described; (3) whether data-driven variable selection was performed; (4) whether methods were used to specifically interpret the contribution of included variables in the prediction. Each item was scored as no (not present), unclear, or yes (present), and then summarized alongside all items to get a study quality score.

## Role of the Funding Source

The funder of the study had no role in study design, data collection, data analysis, data interpretation, or writing of the report. The corresponding author had full access to all the data in the study and had final responsibility for the decision to submit for publication.

## Results

Our database search identified 2728 distinct articles; after a full-text review of 298 papers, 96 were included in the systematic review (Figure 2). Among the included studies, one of the studies used data from a clinical trial, while the others utilized observational data. Of the observational studies, data from cohort studies was the most frequent (34 studies), followed by data from electronic medical records (32 studies), surveys (14 studies) and data from open-access repositories of registry or national survey data (7 studies) (e.g. Scientific Registry of Transplant Recipients Registry^37^). Most of the observational data were structured data (clearly defined data features), while 9 studies included unstructured data (e.g. electrocardiogram, image and heart sound). The earliest year of publication was 1992 (artificial neural network algorithm)^38^, and publications fulfilling our inclusion criteria have been increasing over time (Figure 3). Figure 4 summarizes variables (4A), outcomes (4A), author locations (4B) and types of venues were studies were published (4C). More details on the data sources and populations included, along with all study details are in Table S1.

**Figure 2:**
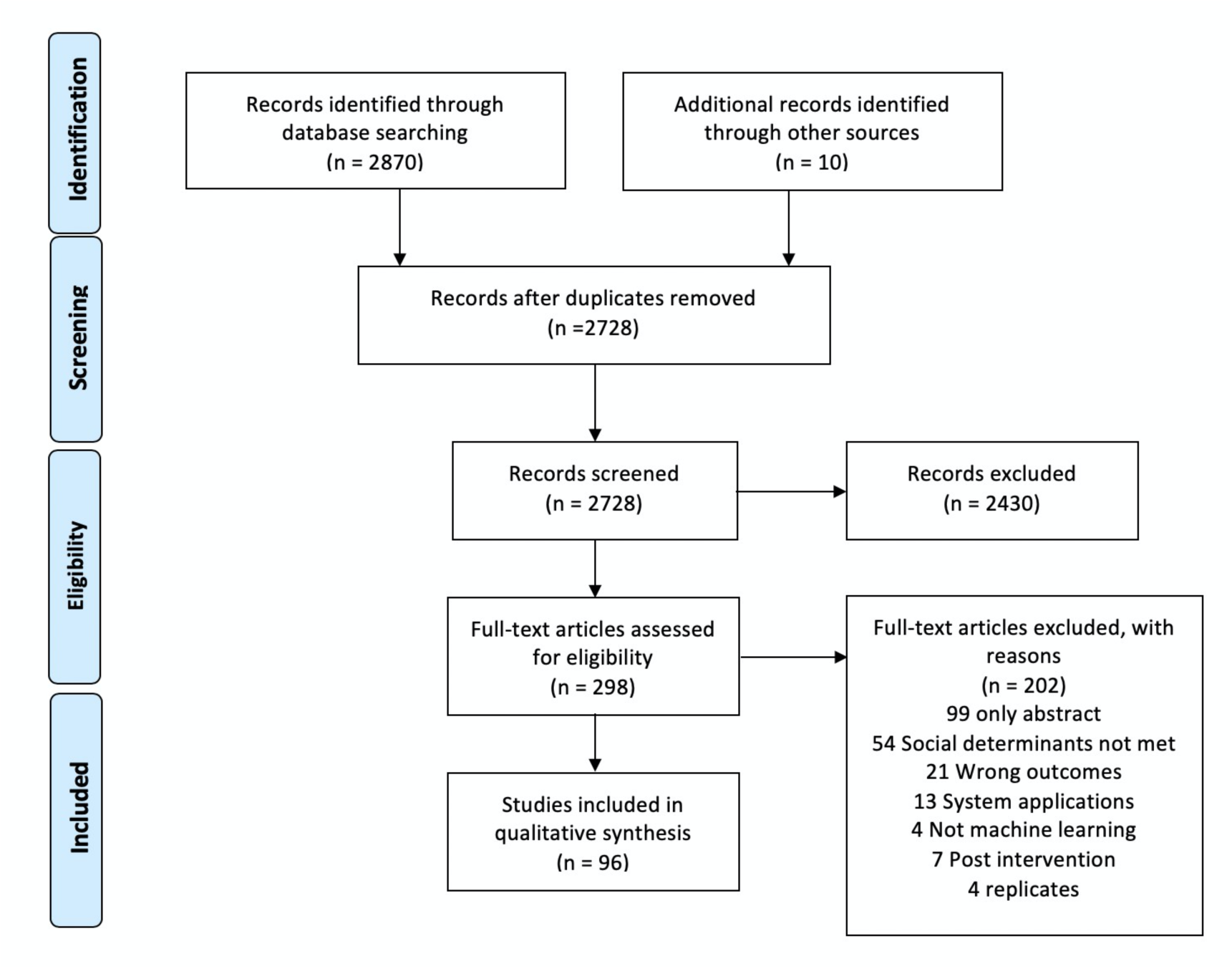
PRISMA flowchart of study review process and exclusion of papers

### Social determinant and variables in the causal pathway

Included studies reported diverse variables across social determinants and variables considered, including race/ethnicity, education, marital status, occupation/employment, individual or household income, medical insurance, area of residence (e.g. urban versus rural or eastern vs. western USA) and other community-level factors of deprivation, income and education and environmental pollutants as well as smoking, alcohol consumption, physical activities, substance abuse and diet. In most studies, gender and age were included as standard variables collected in the survey or EHR. A few studies assessed physical activities and diet as modifiable risk factors for early prevention of CVD.^14,39^ Half of the studies reported feature importance of variables, in which age, gender, smoking (e.g. current smoking/past smoking/non-smoking) and BMI were most frequently reported to contribute significantly to the CVD outcome prediction. Other frequently reported determinants including race/ethnicity, alcohol consumption (e.g. daily intake or alcoholism), and physical activity/exercise (e.g. weekly exercise time). Besides age, gender, BMI and smoking which were frequently reported in all CVD outcomes, alcohol consumption and physical activities were frequently associated with stroke while BMI was frequently associated with coronary artery disease. The top ten variables considered in extracted papers, and their frequency, are illustrated in Figure 4A, which include marital status, education, income and race/ethnicity as the most common social determinants. Four of the studies compared model performance with social determinants and without social determinants; three showed social determinants significantly improved prediction, others showed improved prediction by addition of age, gender and race.^40,41^ The study that showed decreased performance aimed to forecast the pattern of the demand for hemorrhagic stroke healthcare services based on air quality; it is possible that the relationship between specific variable tested and outcome have little direct relationship.^42^

**Figure 3:**
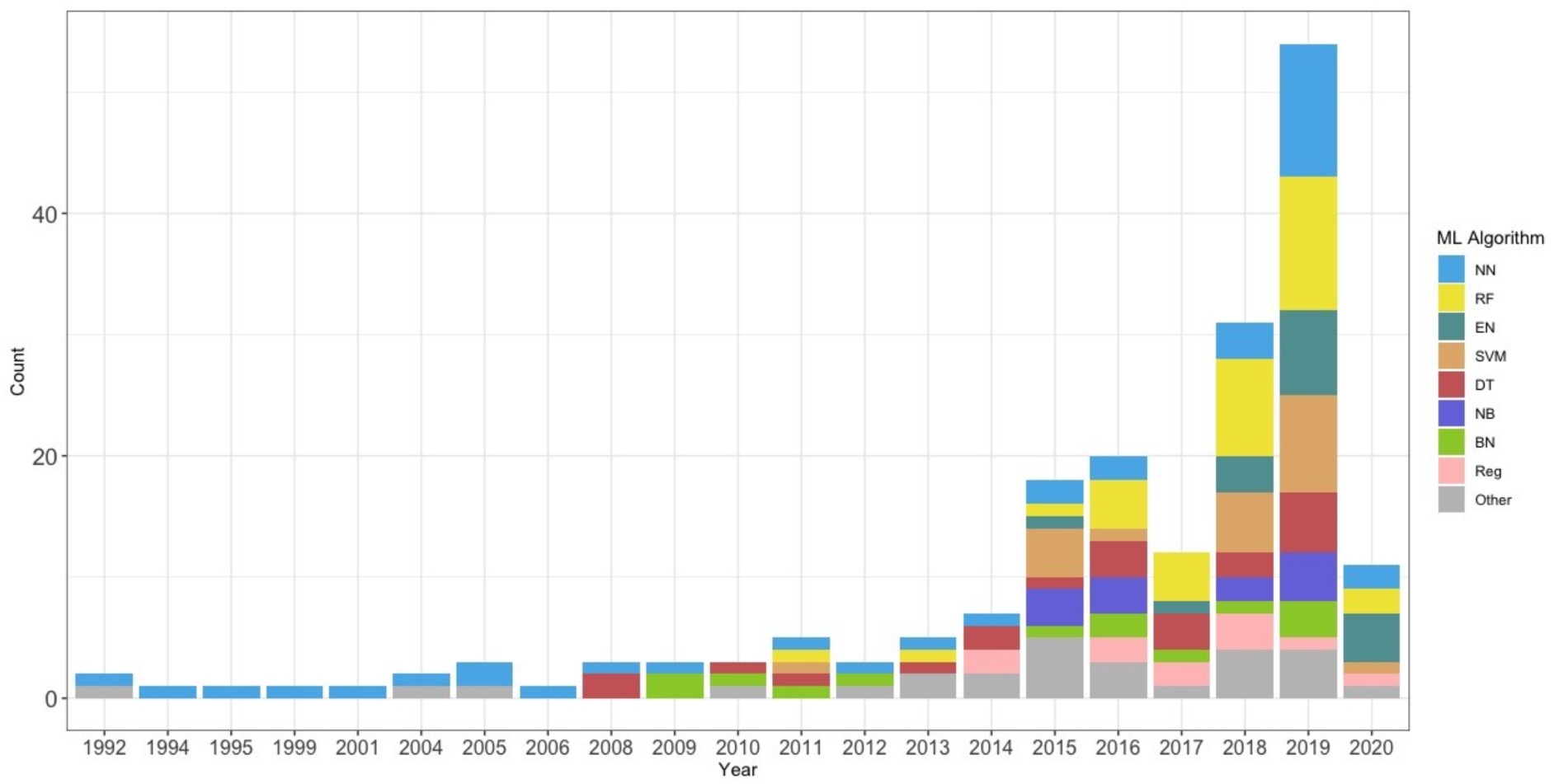
Number of ML algorithms used in publications by year and type (NN: neural network, RF: random forest, EN: ensemble methods (e.g. Adaboost, gradient boosting, bagged decision tree), SVM: support vector machine, DT: decision tree, NB: naïve Bayes, BN: Bayesian network, Reg: regularization methods (ridge/lasso regression), Other: multilayer perceptron, maximum entropy, adversarial network, linear discriminant analysis, k-nearest neighbors, recursive partitioning, clustering, quadratic discriminant, radial basis function kernel)

**Figure 4:**
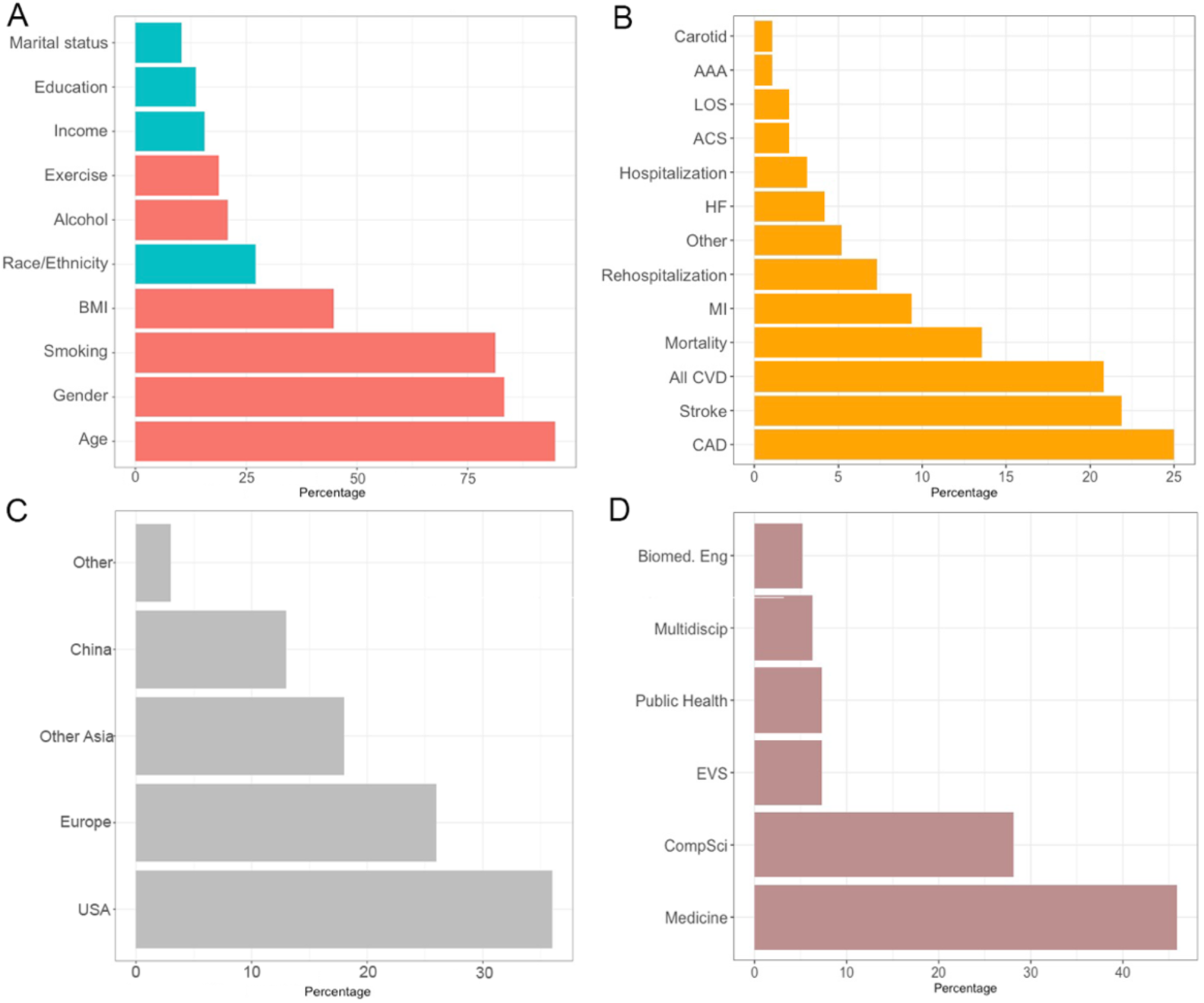
(A) Top ten social determinant and related variables included based on study inclusion criteria (social determinants in blue, other variables in red), (B) most frequently reported CVD outcomes (AAA: abdominal aortic aneurysm, LOS: length of stay, ACS: acute coronary syndrome, HF: heart failure, MI: myocardial infarction, CAD: coronary artery disease) (C) countries of corresponding authors and (D) journal types of publication reported in systematic review papers (EVS: environmental sciences) all with respect to the percentage of included papers they appear in

### Algorithms and model development

The most common machine learning methods were neural network (NN, 36 studies), random forest (RF, 28 studies), and decision trees (DT. 21 studies). Three studies used unsupervised machine learning algorithms, such as clustering to group CVD risk levels or principal component analysis (PCA) to extract features prior to supervised machine learning classification.^14,43,44^ The most frequently used algorithms are described in Table 1. Of the 35 studies using neural networks, 12 used one hidden layer, 23 used multiple hidden layers, including most commonly three-layer perceptron, convolutional neural network and recurrent neural network. Here we refer to these studies collectively as “neural networks” (NN) as deep learning typically refers to an neural network with multiple layers.^45^ Of the 42 studies including multiple machine learning algorithms, random forest (9 studies) and neural network (9 studies) were most frequently reported as the best performing machine learning algorithms. For most commonly studied CVD outcomes, random forest was frequently reported to have the best prediction for stroke while support vector machine (SVM) performed best for coronary artery disease.

**Table 1:** Summary of machine learning algorithms, best performing and sample sizes used in the studies

| Algorithm | Number<br>(%) of<br>papers** | Num as best<br>algorithm when<br>multiple algorithms | Sample Size |  |  |  |
| --- | --- | --- | --- | --- | --- | --- |
|  |  |  | <100 | 100-1000 | 1000-10,000 | >10,000 |
| Neural Net | 35 (36.5%) | 9 | 5 | 8 | 13 | 9 |
| Random Forest | 32 (33.3%) | 9 | 0 | 3 | 14 | 15 |
| Decision Tree | 21 (21.9%) | 2 | 2 | 6 | 8 | 5 |
| Support Vector Machine | 20 (20.8%) | 7 | 1 | 3 | 11 | 5 |
| Ensemble | 17 (17.7%) | 5 | 0 | 2 | 7 | 8 |
| Bayesian Network | 13 (13.5%) | 1 | 1 | 4 | 2 | 6 |
| Naïve Bayes | 12 (12.5%) | 1 | 2 | 2 | 5 | 3 |
| Regularization* | 11 (11.5%) | 0 | 0 | 2 | 6 | 3 |
| Other | 28 (29.2%) | 1 | 4 | 6 | 12 | 6 |

\*regularization included Lasso, Ridge and Elastic net \*\*Note: each paper could include multiple versions or multiple algorithms

There were 24 studies that compared machine learning algorithms with standard linear regression, logistic regression or survival analysis; among those 21 showed improved performance with machine learning. One study of risk prediction for in-hospital mortality in women with ST-elevation myocardial infarction using data from the National Inpatient Sample in the United States, found comparable performance using random forest and logistic regression.^46^ In another study, neural network models for prediction of acute coronary syndromes using clinical data and NN showed similar performance to logistic regression in predicting acute coronary syndrome; however, only 13 variables were considered.^47^ A third study on predicting adverse cardiovascular events by models integrating stress-related ventricular functional and angiographic data showed that while a logistic model demonstrated better performance in this task and implementation, a Bayesian network model showed good performance and also was highlighted as being better at defining causal relationships, and thus useful for designing future models in which new variables can be incorporated in the prediction task.^48^

### Model, validation and performance and study quality

Most studies evaluated the performance of machine learning algorithm(s) developed. Area under the receiver operating characteristic curve (AUC) was the most common evaluation metric used (45) studies, followed by sensitivity (43 studies), specificity (32 studies) and accuracy (32 studies). At least three of the four metrics were used in 31 studies. Other evaluation metrics used included accuracy, positive predictive value, negative predictive value and F1-score, which is the harmonic mean of the precision and recall, commonly used to evaluate machine learning methods via their balance of these metrics. External evaluation was performed in 11 studies, wherein the authors tested the machine learning models developed in one hospital on another hospital or population. For example, one study specifically tested the generalizability of a recurrent neural network model for predicting heart failure risk in a large dataset from 10 hospitals; evaluating the performance of a model trained on each hospital’s training (and validation) sets over the 10 hospitals’ test sets. They also evaluated the model that trained on all hospitals’ training sets over the 10 hospitals’ test sets.^41^ and another used data from one hospital to train neural network models for diagnosis of acute coronary syndrome and tested the model on data from two other hospitals.^47^

Among those reported, most AUC were higher than 0.70 (Figure 5). As most studies were published in biomedical and clinical journals, most studies explicitly interpreted the findings and their relevancy to clinical applications. Almost half (40/96) of the studies compared more than one machine learning algorithm, of which Random Forest was most commonly the best performing model. The mean score of included studies in the 4-item quality assessment scale (based on evaluation of ML, data-driven selection of features, hyperparameter tuning description, interpretation of the model) was 3.34. Half of the studies (49) had full scores and 30 studies missed one of the four items. Commonly missed items were data-driven feature selection and details of hyperparameter tuning (cross-validation or grid search strategies were utilized in 68 studies to tune hyperparameters; other studies didn’t give details about hyper-parameter tuning process). Half of all the studies utilized a data-driven selection method to identify features before fitting machine learning models, which is defined as extracting a subset of useful variables among the original variables and transforming data from a high- to a low-dimensional space.^49^ As deep learning models to extract features while training, those studies did not always include a feature selection process.

**Figure 5:**
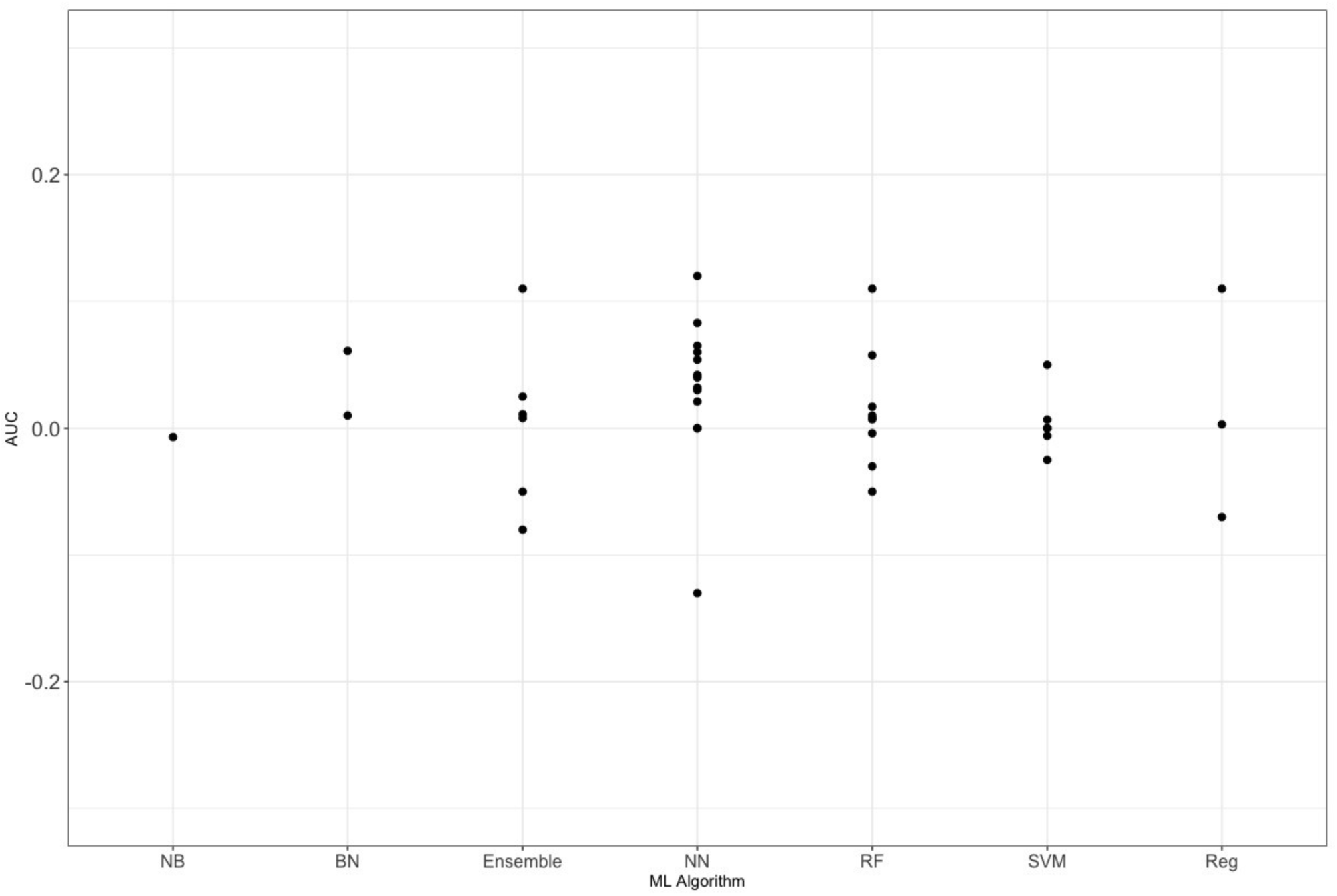
Difference of Area under the ROC curve between ML and LR by ML algorithm type

## Discussion

To our knowledge, this is the first systematic review to illustrate how machine learning is being used to integrate SDoH in cardiovascular disease prediction models. This review distills which types of algorithms and SDoH and related variables have been considered and resulting performance. We found that the flexibility of machine learning models has proved useful in CVD prediction models, with them commonly performing better than regression approaches. We find that models that consider SDoH and related variables also benefit from flexible modeling approaches, with neural networks consistently outperforming regression across all CVD outcomes.

Broadly, we found several limitations in the content covered by included papers. First, the studies were highly skewed to originate from USA, Europe and China, with lower-income locations not being well represented. Moreover, we found that the race and ethnicity distribution in some studies was also not very representative of underlying populations. This is particularly striking given the high and increasing CVD burden and changing socio-environmental circumstances in lower-income countries and regions and disparities in CVD burden. The variance of social determinants incorporated into models, and thus the performance and applicability of those models across contexts, will be decreased with less diversity in the study sample.^50^ SDoH variables themselves were also not very frequently included, with only marital status, education, income and race/ethnicity in the top ten. Environmental attributes that have been shown as important modifiable components of CVD risk such as green spaces and stress^32,51,52^ were very few, and even then were very broad (e.g. region of country).^42^ If such area-based variables are included, machine learning may also prove useful to unweave the strands of environmental influences but also integrate the effects of the various components of the environment into a comprehensive model.^32^ We also find that models did not take into account social processes associated with socioeconomic conditions across the life course. Socioeconomic position, psychosocial factors and behaviors during adolescence and youth are important likely to be important in the development of CVD and precursors (dyslipidemia, hypertension, and smoking).^53^ Finally, studies generally included gender interchangeably with sex, which precludes consideration of the socially-determined aspect of gender.^54^

Despite these limitations, our results largely found social determinants and variables considered to improve model performance. In terms of algorithms, several types of machine learning algorithms were evaluated, with results showing that when compared within studies, the most flexible models such as neural networks and random forest models were best performing. Neural networks also most commonly outperformed regression models. This is understood to be the case because neural networks include hidden layers which can take into account more complex relations in the data, and therefore this may be another possible explanation for the improved performance.^55,56^ Moreover, recent studies uncovering network and spillover effects (social environment) and shared decision-making^57^ involved in physical activity,^58,59^ diet^60^ and smoking^61^ indicate that the pathways that inform these behaviors are intricate. However, this may illuminate an opportunity for machine learning, which based on flexibility, can help capture such complex interactions.

The constraints on included data are likely due to difficulties in capturing certain SDoH variables and linking them with individual records in databases used in many of the included studies. Studies have largely used social variables from available data sources; commonly those in the electronic health record. The use of flexible, machine learning models also bring concerns regarding interpretability and potential over-fitting to data,^62^ though this was not a common discussion topic across all papers. This is likely because most models selected variables based on prior clinical significance, thus prediction performance would be based on such factors which are known to be relevant to CVD even if the specific importance of each variable was not measured. Furthermore, most papers (66) papers used methods such as automatic relevance determination^55^ or feature selection^63^ to examine and/or rank the importance of variables in machine learning models. This was the case even as articles were published in a variety of venues (Figure 4D).

While this is the first review that gives findings related to the use of machine learning and social determinants for CVD prediction, there are individual studies that support components of the findings of this study. First, machine learning in general has shown promise with respect to cardiovascular disease prediction.^64-66^ Compared to the established American College of Cardiology/American Heart Association risk calculator to predict incidence and prognosis of ASCVD,^20^ previous work has shown that machine-learning algorithms (especially random forest, gradient boosting machines and neural networks) were better at identifying individuals who will develop CVD and those who will not.^17-19^ These studies have attributed this to the fact that standard CVD risk assessment models make an implicit assumption that each risk factor is related in a linear fashion to CVD outcomes and such models may thus oversimplify complex relationships which include large numbers of risk factors with non-linear interactions. The role of social determinants in cardiovascular disease (not specifically machine learning-related) has been studied through several papers and systematic reviews. While full summaries of this work have been performed elsewhere,^11^ we note that there have been several studies of various proximal and distal social determinants and cardiovascular disease. In general, studies indicate that the changing burden of disease due to societal and environmental conditions, as well as increasing advances in treatment and prevention have not been shared equally across economic, racial and ethnic groups, compelling the need for broad range consideration of social determinants in CVD prediction.^11,31^ Finally, the models that have incorporated social determinants and machine learning for CVD prediction also reflect limitations of many machine learning algorithms that have been highlighted recently, which are based on homogenous populations, particular with respect to race (captured through the limited geographic diversity in Figure 4C).^17^

Our review was limited in several aspects. First, the included studies evaluated different types of cardiovascular outcomes, and heterogeneity of outcome metrics makes it difficult to compare machine learning performance across studies. The population considered also includes samples from different data sources, hospitals and countries which taken together make the comparison across studies not standardized. Third, most studies did not evaluate external validity, leaving the applicability of the algorithms to other populations or healthcare settings inconclusive. Fourth, the review was also limited to studies published in English, which might have created some bias in the articles that were ultimately retained for the analysis.

Findings emphasize the need to comprehensively capture both proximal and distal social determinant variables in models. Where mechanisms are not well understood, machine learning can also be used to understand relationships between social and biological variables comprehensively. For example, race is often conceptualized as a proxy for variables for socioeconomic position or cultural factors and better ways to capture as well as understand relationships between these factors and their impact on CVD risk should be investigated. Indeed, identification of potential mediating and moderating factors in these pathways of social determinants will inform public health interventions. Improved constructs will also help in incorporation of environmental and behavioral variables such as diet and physical activity which were not well represented in current studies. Our findings support bodies of work that promote inclusion of such information in the electronic health record^67,68^ and we add reasoning that this would also enable study of social determinants in machine learning in large enough sample sizes to reduce overfitting of models. Finally, results emphasize the need for studies that include more diverse populations with varied environmental and social influences, which would represent and ensure validity of prediction models across these diverse interactions^50^ to improve cardiovascular disease prediction in diverse settings, in particular those where disease risk is increasing.

## Data Availability

All papers included in the review are summarized in the Appendix.

## Research in context

### Evidence before this study

While there are no reviews that specifically address social determinants, machine learning and cardiovascular disease (CVD), the latest research on cardiovascular disease indicates the imperative relevance of social determinants. Societal and environmental conditions distributed unequally among groups are driving a significant and increasing global burden of cardiovascular diseases particularly to low and middle-income countries as well as lower-socioeconomic groups in high-income countries. At the same time, research shows that machine learning offers the potential for capturing flexible relationships compared to the linear relationships assumed in typical CVD risk scores, which is of particular relevance for the consideration of social variables. Select models have examined the performance of certain social variables in CVD prediction, including those that use machine learning, but given these rapidly advancing research areas, a systematic examination into which social determinants have been modelled and how different methods have performed, is needed.

### Added value of this study

Through a rigorous and comprehensive systematic review, we assessed the state-of-the art methods prediction of the two types of CVD with the highest recent mortality (ischemic heart disease and stroke), that use machine learning and incorporate social determinants and related variables in their causal pathway. We show that environmental and area-based determinants are lacking from most models. Machine learning, especially flexible models such as neural networks, show good performance in relation to regression models. We accounted for model and information gain differences across by examining within study performance of best algorithm compared to regression. We assessed the quality of their implementations via best-practices from the machine learning literature, finding that quality was generally rigorous. Finally, we show that the origin of studies is highly skewed to USA and middle/high-income countries in Europe and Asia, which indicates that knowledge regarding the diversity of social determinants and their impact is limited.

### Implications of all the available evidence

With the significant burden of CVD and large burden in low- and middle-income countries, this work directly informs how we can augment prediction models, using state of the art machine learning methods, while also taking into account growing social, environmental risk factors that shape CVD risk. According to the findings of this review, strategies to capture social variables, especially environmental determinants are needed in the electronic health record databases from which machine learning methods are commonly developed. Finally, studies to-date represent a narrow set of locations; we need to support studies in low- and middle-income countries to identify and tailor our understanding to the specific social determinants in these populations.

## Acknowledgements

We thank Dorice Vieira for valuable help with the search process.

## Appendix

**Table S1:**
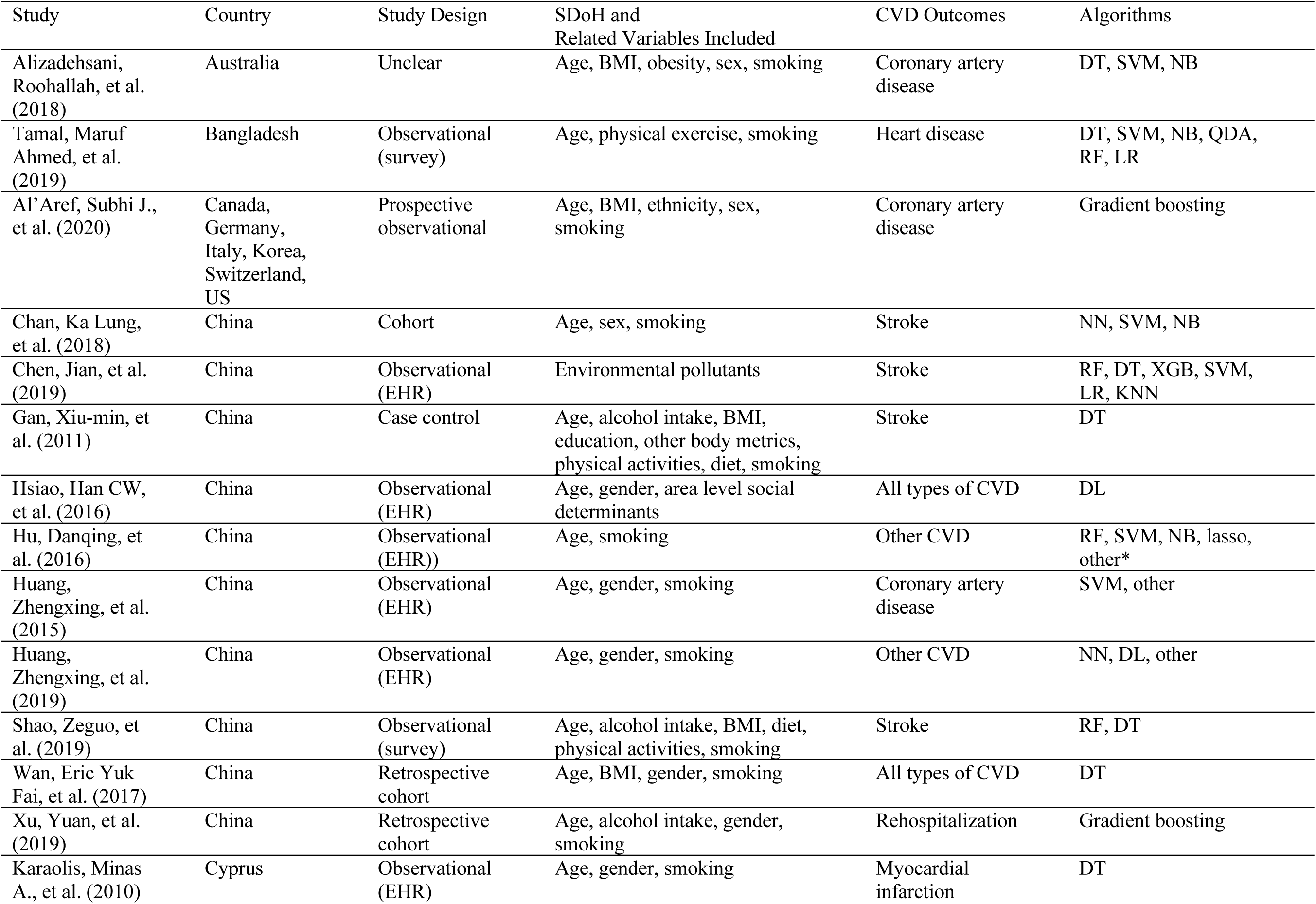
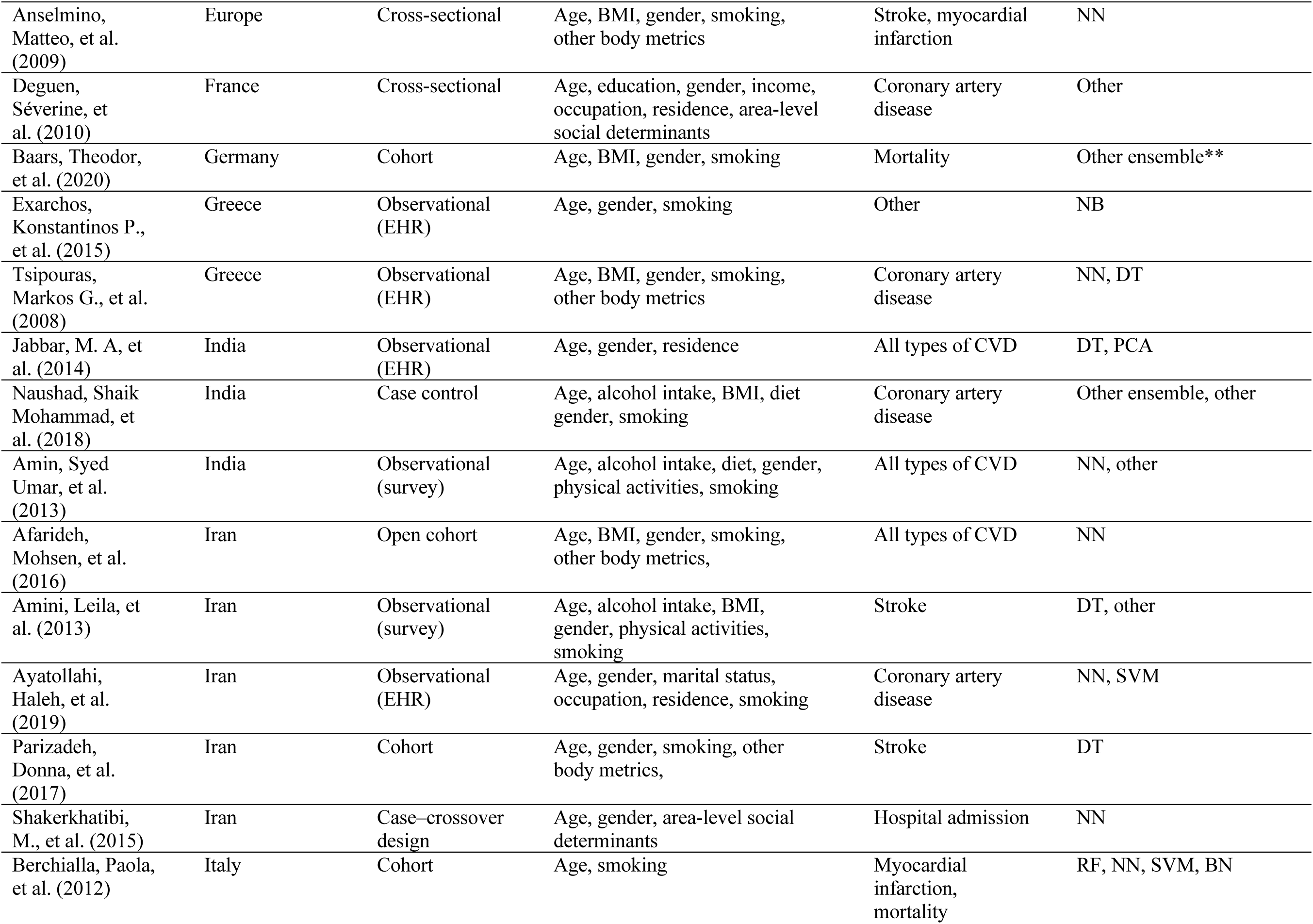
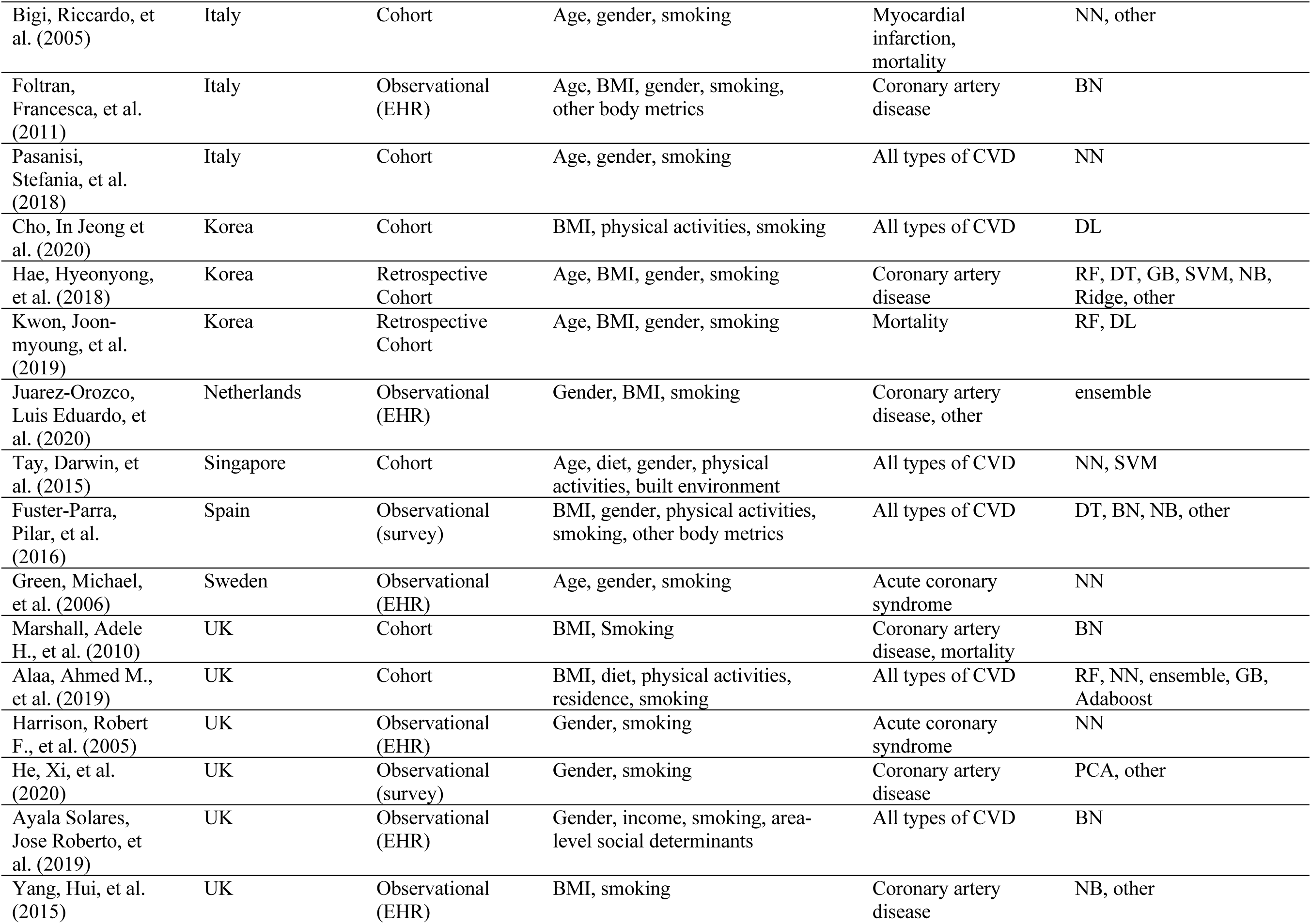
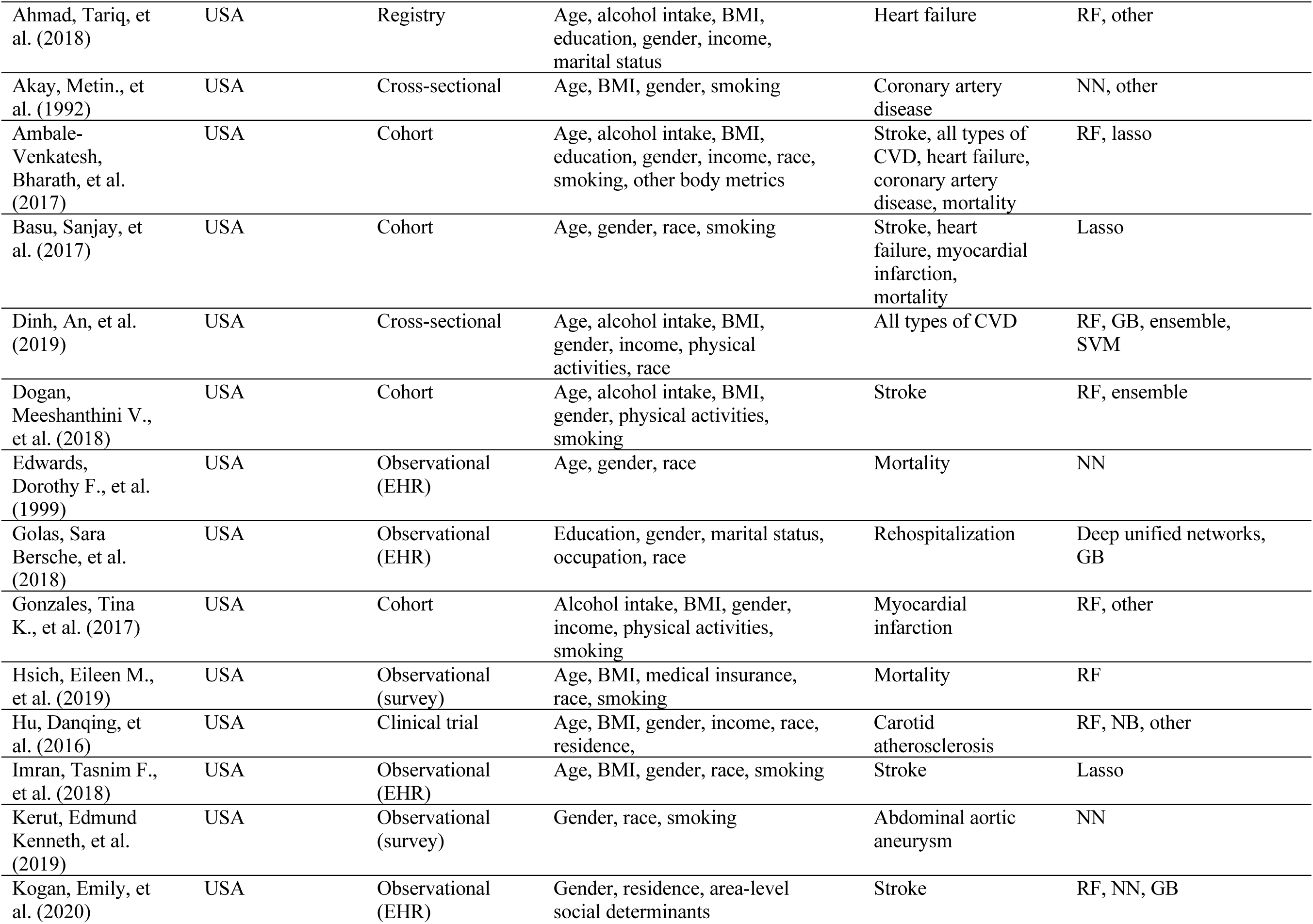
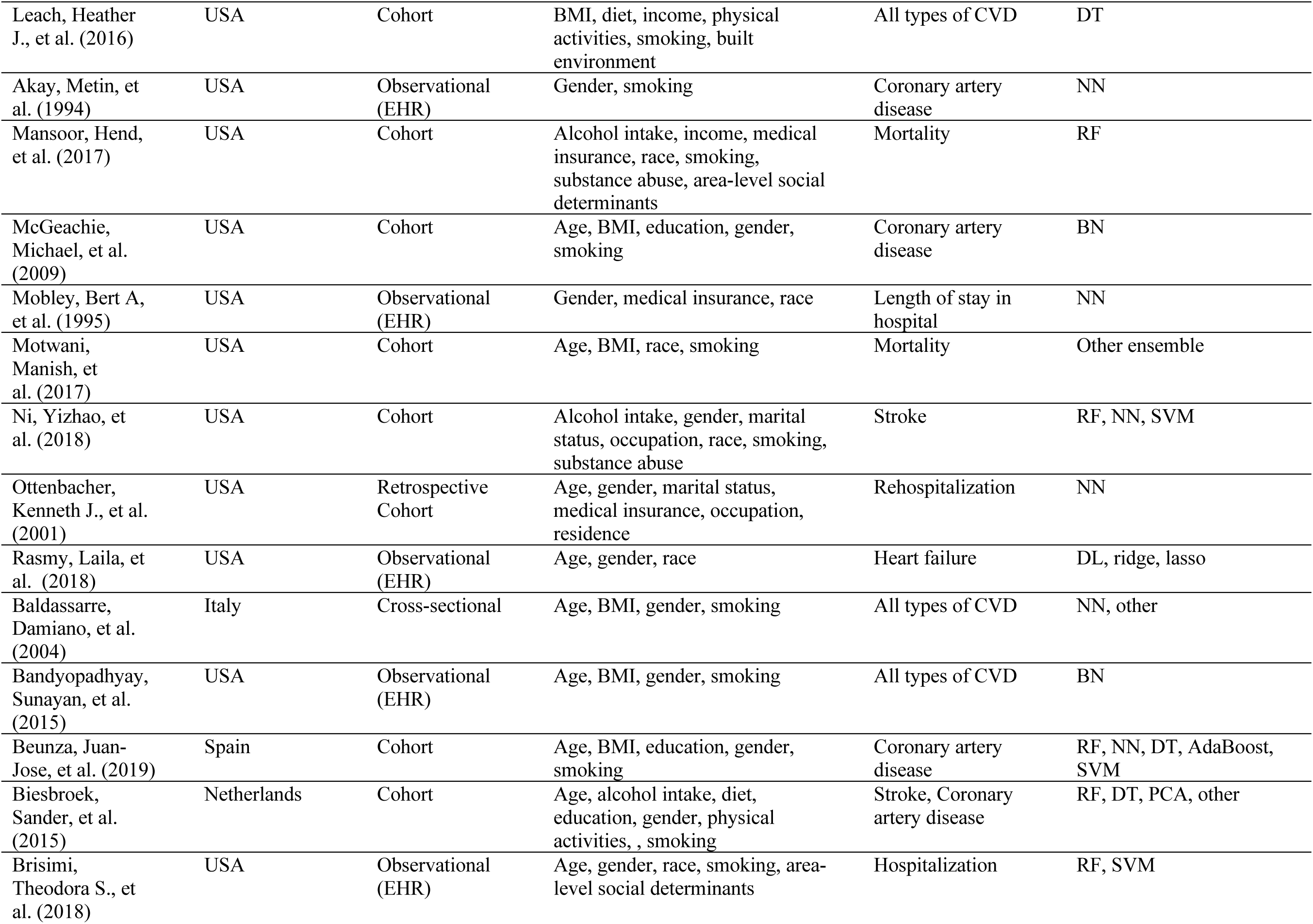
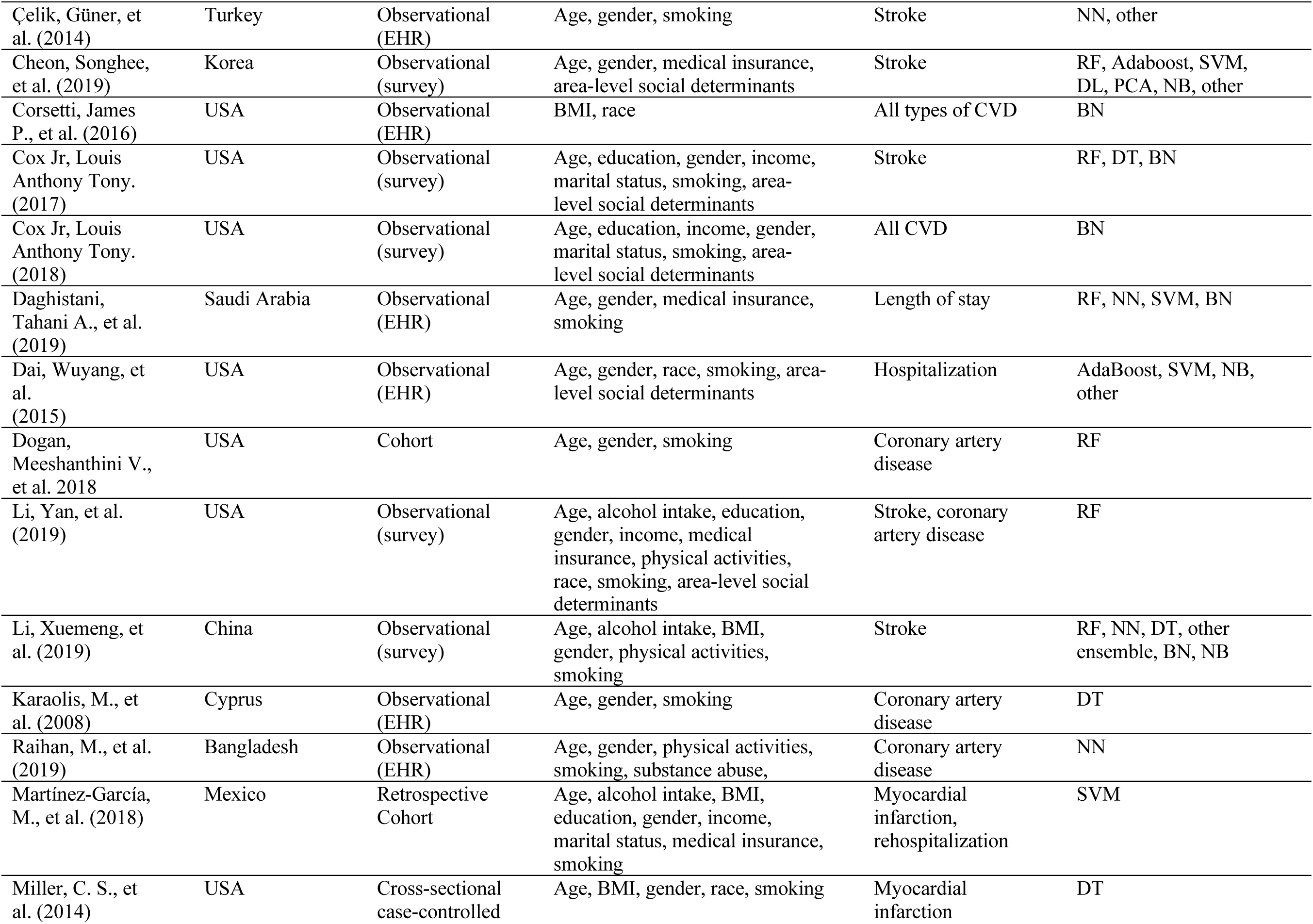
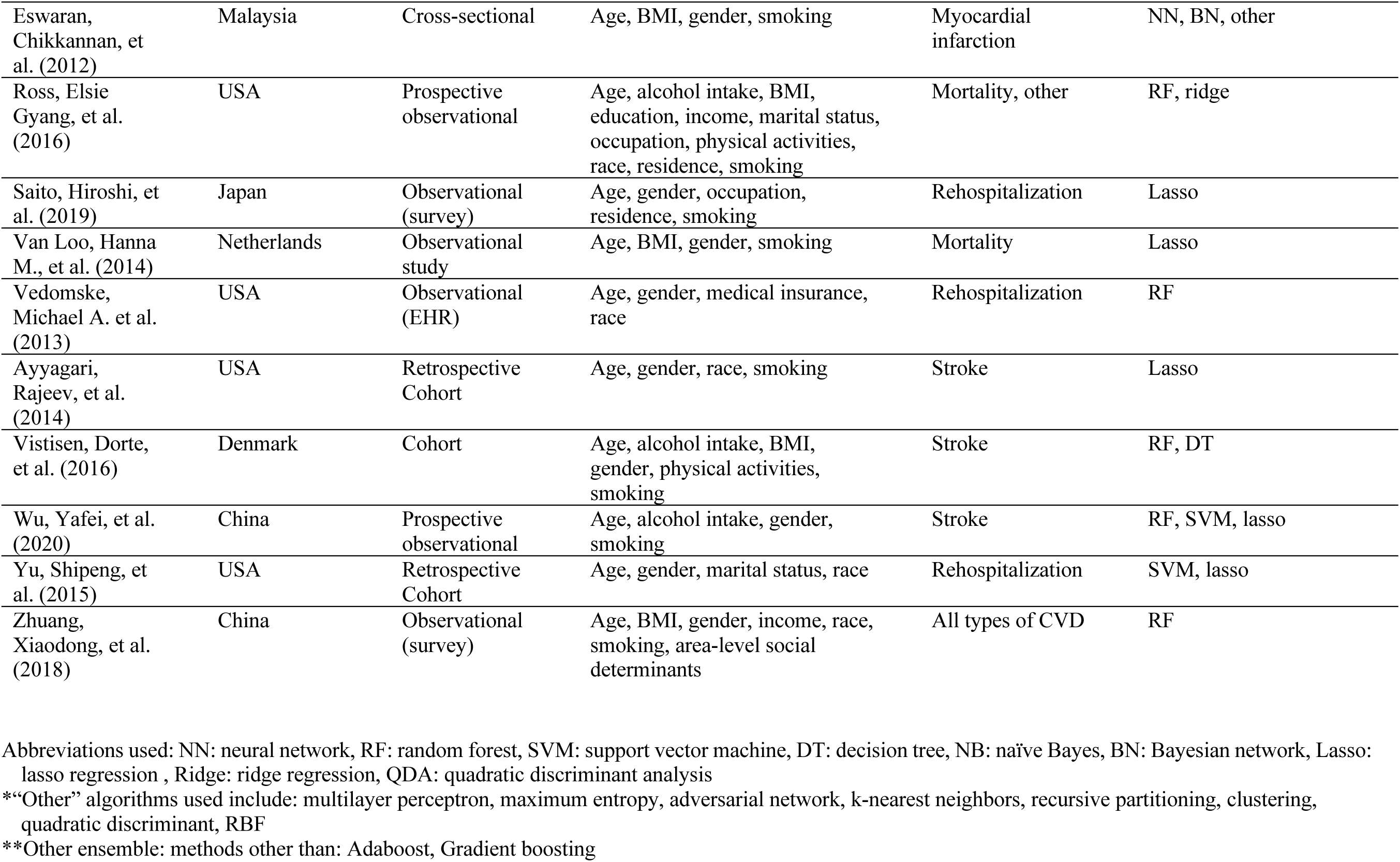
Summary of all included papers (attached at end).

### Search terms and search strategies PubMed

(Social determinants of health[Mesh] OR demography[Mesh] OR demographic*[tw] OR race [tw] OR racism[Mesh] OR “ethnicity”[tw] OR gender identity[Mesh] OR gender[tw] OR social[tw] OR social support[Mesh] OR income[Mesh] OR education[Mesh] OR employment[Mesh] OR marital status[Mesh] OR occupation[tw] OR “health insurance”[tw] OR health literacy[Mesh] OR marriage[tw] OR insurance[tw] OR housing[tw] OR home[tw] OR religion[tw] OR socioeconomic factors[Mesh] OR social class[Mesh] OR “social status”[tw] OR “access healthcare”[tw] OR healthcare disparities[Mesh] OR “financial difficulties”[tw] OR poverty[Mesh] OR “social disparity”[tw] OR unemployment[Mesh] OR social condition[Mesh] OR “social inequality”[tw] OR vulnerable population[Mesh] OR “social environment”[tw] OR sociodemographic*[tw] OR sociological factors[Mesh] OR body mass index[Mesh] OR physical activity[Mesh] OR diet[Mesh] OR smoking[Mesh] OR “alcohol consumption”[tw] OR tobacco[Mesh] OR “substance use”[tw] OR “physical inactivity”[tw] OR “substance abuse”[tw] OR health* behavi*r*[tw] OR health* service[tw] OR environment[Mesh] OR “living environment” OR “birthplace”[tw] OR “pollution”[tw] OR residence characteristics[Mesh] OR “geographic locations”[tw] OR “rural”[tw] OR “urban health”[tw] OR neighborhood[tw] OR cultur*[tw]) AND (machine learning[Mesh] OR supervised machine learning[Mesh] OR decision trees[Mesh] OR neural networks[Mesh] OR “Naive Bayes”[tw] OR “kNN”[tw] OR support vector machine[Mesh] OR perceptron[tw] OR “radial basis function”[tw] OR “Bayesian Network”[tw] OR “random forest”[tw] OR “classification tree”[tw] OR “elastic net”[tw] OR “multilayer perceptron”[tw] OR lasso[tw] OR ridge[tw] OR “nearest neighbor”[tw] OR deep learning[Mesh] OR boosting[tw] OR bagging[tw] OR ensemble[tw]) AND (“atherosclerotic cardiovascular disease”[tw] OR cardiovascular abnormalities[Mesh] OR heart disease*[tw] OR heart arrest[Mesh] OR myocardial ischemia[Mesh] OR arterial occlusive diseases[Mesh] OR cerebrovascular disorders[Mesh] OR peripheral vascular diseases[Mesh]) **Embase:** (exp “social determinants of health”/ or exp ’’demography”/ or demographic* or *race”/ or ’’racism” or *ethnicity”/ or exp “gender identity”/ or “gender” or “social” or exp “social support”/ or exp “education”/ or exp “employment”/ or “income” or “marital status” or exp “occupation”/ or exp “health insurance”/ or “health literacy”/ or exp “marriage”/ or “insurance” or exp “housing”/ or “home” or “religion” or “socioeconomic factors” or exp “socioeconomics”/ or “social class” or exp “healthcare access”/ or exp “health care disparities”/ or “financial difficulties” or exp “poverty”/ or “social disparity” or exp “unemployment”/ or exp “social status”/ or “social inequality” or exp “vulnerable population”/ or *social environment/ or sociodemographic* or *body mass/ or *physical activity/ or *diet/ or exp “smoking”/ or exp “alcohol consumption”/ or “tobacco use” or exp “substance use”/ or exp “physical inactivity”/ or exp “substance abuse”/ or *environment/ or *birthplace/ or exp “pollution”/ or “residence characteristics” or *geography/ or “neighborhood” or cultur* or exp “rural health”/ or exp “urban health”/) and (exp “machine learning”/ or “supervised machine learning” or exp “decision trees”/ or “neural networks” or exp “artificial neural network”/ or “Naive Bayes” or exp “Bayesian learning”/ or exp “k nearest neighbor”/ or “knn” or exp “support vector machine”/ or “SVM” or exp “perceptron”/ or exp “radial based function”/ or “Bayesian Network” or exp “random forest”/ or “classification tree” or “elastic net” or “multilayer perceptron” or “lasso” or “ridge” or exp “deep learning”/ or “boosting” or “ensemble”) and (”atherosclerotic cardiovascular disease” or *coronary artery atherosclerosis/ or “cardiovascular abnormalities” or exp “cardiovascular malformation”/ or *heart disease/ or exp “heart arrest”/ or exp “myocardial ischemia”/ or “arterial occlusive diseases” or exp “cerebrovascular disorders”/ or exp “peripheral vascular diseases”/)

### Web of Science

TS=((“Social determinants of health” OR demography OR demographic* OR race OR ethnicity OR “gender identity” OR gender OR social OR “social support” OR income OR education OR employment OR “marital status” OR occupation OR “health insurance” OR marriage OR insurance OR housing OR religion OR “socioeconomic factors” OR “social class” OR “access healthcare” OR “healthcare disparities” OR “financial difficult” OR poverty OR “social disparity” OR unemployment OR “social condition” OR “social inequality” OR “vulnerable population” OR “social environment” OR sociodemographic* OR “body mass index” OR “physical activity” OR diet OR smoking OR “alcohol consumption” OR tobacco OR “substance use” OR “physical inactivity” OR “substance abuse” OR environment OR birthplace OR pollution OR “residence characteristics” OR “geographic locations” OR “rural” OR “urban health”) AND (“machine learning” OR “supervised machine learning” OR “decision trees” OR “neural networks” OR “Naive Bayes” OR kNN OR “support vector machine” OR “perceptron” OR “radial basis function” OR “Bayesian Network” OR “random forest” OR “classification tree” OR “elastic net” OR “multilayer perceptron” OR “lasso” OR “ridge” OR “nearest neighbor” OR “deep learning” OR “boosting” OR “ensemble”) AND (“atherosclerotic cardiovascular disease” OR “cardiovascular abnormalities” OR heart disease* OR “heart arrest” OR “myocardial ischemia” OR “arterial occlusive diseases” OR “cerebrovascular disorders” OR “peripheral vascular diseases”))

### IEEE

(“Social determinants of health” OR demography OR demographic* OR race OR ethnicity OR “gender identity” OR gender OR social OR “social support” OR income OR education OR employment OR “marital status” OR occupation OR “health insurance” OR marriage OR insurance OR housing OR religion OR “socioeconomic factors” OR “social class” OR “access healthcare” OR “healthcare disparities” OR “financial difficult” OR poverty OR “social disparity” OR unemployment OR “social condition” OR “social inequality” OR “vulnerable population” OR “social environment” OR sociodemographic* OR “body mass index” OR “physical activity” OR diet OR smoking OR “alcohol consumption” OR tobacco OR “substance use” OR “physical inactivity” OR “substance abuse” OR environment OR birthplace OR pollution OR “residence characteristics” OR “geographic locations” OR “rural” OR “urban health”) AND (“machine learning” OR “supervised machine learning” OR “decision trees” OR “neural networks” OR “Naive Bayes” OR kNN OR “support vector machine” OR “perceptron” OR “radial basis function” OR “Bayesian Network” OR “random forest” OR “classification tree” OR “elastic net” OR “multilayer perceptron” OR “lasso” OR “ridge” OR “nearest neighbor” OR “deep learning” OR “boosting” OR “ensemble”) AND (“atherosclerotic cardiovascular disease” OR “cardiovascular abnormalities” OR heart disease* OR “heart arrest” OR “myocardial ischemia” OR “arterial occlusive diseases” OR “cerebrovascular disorders” OR “peripheral vascular diseases”)

## Notes

### Competing Interest Statement

The authors have declared no competing interest.

### Author Declarations

N/A (Systematic review)

## References

1 World Health Organization. Cardiovascular diseases (CVDs) fact sheet. World Health Organization (2017).

2 Deaton, C. et al. The global burden of cardiovascular disease. European Journal of Cardiovascular Nursing 10, S5-S13 (2011).

3 Heidenreich, P. A. et al. Forecasting the impact of heart failure in the United States: a policy statement from the American Heart Association. Circulation: Heart Failure 6, 606–619 (2013).

4 Kuzawa, C. W. & Sweet, E. Epigenetics and the embodiment of race: developmental origins of US racial disparities in cardiovascular health. American Journal of Human Biology: The Official Journal of the Human Biology Association 21, 2–15 (2009).

5 Carnethon, M. R. et al. Cardiovascular health in African Americans: a scientific statement from the American Heart Association. Circulation 136, e393-e423 (2017).

6 Stuckler, D., McKee, M., Ebrahim, S. & Basu, S. Manufacturing epidemics: the role of global producers in increased consumption of unhealthy commodities including processed foods, alcohol, and tobacco. PLoS Med 9, e1001235 (2012).

7 Lakka, T. A. et al. Sedentary lifestyle, poor cardiorespiratory fitness, and the metabolic syndrome. Medicine & Science in Sports & Exercise (2003).

8 Health, W. C. o. S. D. o. & Organization, W. H. Closing the gap in a generation: health equity through action on the social determinants of health: Commission on Social Determinants of Health final report. (World Health Organization, 2008).

9 Joseph, P. et al. Reducing the global burden of cardiovascular disease, part 1: the epidemiology and risk factors. Circulation research 121, 677–694 (2017).

10 Tillmann, T. et al. Psychosocial and socioeconomic determinants of cardiovascular mortality in Eastern Europe: A multicentre prospective cohort study. PLoS medicine 14, e1002459 (2017).

11 Havranek, E. P. et al. Social determinants of risk and outcomes for cardiovascular disease: a scientific statement from the American Heart Association. Circulation 132, 873–898 (2015).

12 Theodore, R. F. et al. Childhood to early-midlife systolic blood pressure trajectories: early-life predictors, effect modifiers, and adult cardiovascular outcomes. Hypertension 66, 1108–1115 (2015).

13 Cooper, R. et al. Trends and disparities in coronary heart disease, stroke, and other cardiovascular diseases in the United States: findings of the national conference on cardiovascular disease prevention. Circulation 102, 3137–3147 (2000).

14 He, X., Matam, B. R., Bellary, S., Ghosh, G. & Chattopadhyay, A. K. cHD Risk Minimization through Lifestyle control: Machine Learning Gateway. Scientific reports 10, 1–10 (2020).

15 Watson, D. S. et al. Clinical applications of machine learning algorithms: beyond the black box. Bmj 364 (2019).

16 Rajkomar, A., Dean, J. & Kohane, I. Machine learning in medicine. New England Journal of Medicine 380, 1347–1358 (2019).

17 Alaa, A. M., Bolton, T., Di Angelantonio, E., Rudd, J. H. & van der Schaar, M. Cardiovascular disease risk prediction using automated machine learning: A prospective study of 423,604 UK Biobank participants. PloS one 14, e0213653 (2019).

18 Dimopoulos, A. C. et al. Machine learning methodologies versus cardiovascular risk scores, in predicting disease risk. BMC Medical Research Methodology 18, 179 (2018).

19 Kakadiaris, I. A. et al. Machine learning outperforms ACC/AHA CVD risk calculator in MESA. Journal of the American Heart Association 7, e009476 (2018).

20 Cook, N. R. & Ridker, P. M. Further insight into the cardiovascular risk calculator: the roles of statins, revascularizations, and underascertainment in the Women’s Health Study. JAMA internal medicine 174, 1964–1971 (2014).

21 Caballero, F. F. et al. Advanced analytical methodologies for measuring healthy ageing and its determinants, using factor analysis and machine learning techniques: the ATHLOS project. Scientific Reports 7, 43955 (2017).

22 Seligman, B., Tuljapurkar, S. & Rehkopf, D. Machine learning approaches to the social determinants of health in the health and retirement study. SSM-population health 4, 95–99 (2018).

23 People, C. o. L. H. I. f. H., Health, B. o. P., Practice, P. H. & Medicine, I. o. Leading health indicators for healthy people 2020: letter report. (National Academies Press, 2011).

24 Council, N. R. & Population, C. o. US health in international perspective: Shorter lives, poorer health. (National Academies Press, 2013).

25 Short, S. E. & Mollborn, S. Social determinants and health behaviors: conceptual frames and empirical advances. Current opinion in psychology 5, 78–84 (2015).

26 Shiroma, E. J. & Lee, I.-M. Physical activity and cardiovascular health: lessons learned from epidemiological studies across age, gender, and race/ethnicity. Circulation 122, 743–752 (2010).

27 Nuttall, F. Q. Body mass index: obesity, BMI, and health: a critical review. Nutrition today 50, 117 (2015).

28 Kotsiantis, S. B., Zaharakis, I. & Pintelas, P. Supervised machine learning: A review of classification techniques. Emerging artificial intelligence applications in computer engineering 160, 3–24 (2007).

29 LeCun, Y., Bengio, Y. & Hinton, G. Deep learning. nature 521, 436–444 (2015).

30 Roth, G. A. et al. Global, regional, and national age-sex-specific mortality for 282 causes of death in 195 countries and territories, 1980-2017: a systematic analysis for the Global Burden of Disease Study 2017. The Lancet 392, 1736–1788 (2018).

31 Kreatsoulas, C. & Anand, S. S. The impact of social determinants on cardiovascular disease. Canadian Journal of Cardiology 26, 8C-13C (2010).

32 Bhatnagar, A. Environmental determinants of cardiovascular disease. Circulation research 121, 162–180 (2017).

33 Cheng, I., Ho, W. E., Woo, B. K. & Tsiang, J. T. Correlations between health insurance status and risk factors for cardiovascular disease in the elderly Asian American population. Cureus 10 (2018).

34 Fang, J. et al. Association of birthplace and coronary heart disease and stroke among US adults: National Health Interview Survey, 2006 to 2014. Journal of the American Heart Association 7, e008153 (2018).

35 Lapane, K. L., Lasater, T. M., Allan, C. & Carleton, R. A. Religion and cardiovascular disease risk. Journal of Religion and Health 36, 155–164 (1997).

36 Chen, P.-H. C., Liu, Y. & Peng, L. How to develop machine learning models for healthcare. Nature materials 18, 410 (2019).

37 Hsich, E. M. et al. Variables of importance in the Scientific Registry of Transplant Recipients database predictive of heart transplant waitlist mortality. American Journal of Transplantation 19, 2067–2076 (2019).

38 Akay, M. Noninvasive diagnosis of coronary artery disease using a neural network algorithm. Biological cybernetics 67, 361–367 (1992).

39 Shao, Z., Chen, C., Li, W., Ren, H. & Chen, W. Assessment of the risk factors in the daily life of stroke patients based on an optimized decision tree. Technology and Health Care 27, 317–329 (2019).

40 McGeachie, M. et al. An integrative predictive model of coronary artery calcification in arteriosclerosis. Circulation 120, 2448 (2009).

41 Rasmy, L. et al. A study of generalizability of recurrent neural network-based predictive models for heart failure onset risk using a large and heterogeneous EHR data set. Journal of biomedical informatics 84, 1116 (2018).

42 Chen, J. et al. Machine Learning-Based Forecast of Hemorrhagic Stroke Healthcare Service Demand considering Air Pollution. Journal of healthcare engineering 2019 (2019).

43 Cheon, S., Kim, J. & Lim, J. The use of deep learning to predict stroke patient mortality. International journal of environmental research and public health 16, 1876 (2019).

44 Jabbar, M., Deekshatulu, B. & Chndra, P. in International Conference on Circuits, Communication, Control and Computing. 322-328 (IEEE).

45 Illing, B., Gerstner, W. & Brea, J. Biologically plausible deep learning—But how far can we go with shallow networks? Neural Networks 118, 90–101 (2019).

46 Mansoor, H., Elgendy, I. Y., Segal, R., Bavry, A. A. & Bian, J. Risk prediction model for in-hospital mortality in women with ST-elevation myocardial infarction: A machine learning approach. Heart & Lung 46, 405411 (2017).

47 Harrison, R. F. & Kennedy, R. L. Artificial neural network models for prediction of acute coronary syndromes using clinical data from the time of presentation. Annals of emergency medicine 46, 431–439 (2005).

48 Berchialla, P., Foltran, F., Bigi, R. & Gregori, D. Integrating stress-related ventricular functional and angiographic data in preventive cardiology: a unified approach implementing a Bayesian network. Journal of Evaluation in Clinical Practice 18, 637–643 (2012).

49 Chu, C. et al. Does feature selection improve classification accuracy? Impact of sample size and feature selection on classification using anatomical magnetic resonance images. Neuroimage 60, 59–70 (2012).

50 Harper, S., Lynch, J. & Smith, G. D. Social determinants and the decline of cardiovascular diseases: understanding the links. Annual review of public health 32, 39–69 (2011).

51 Richardson, E. A. & Mitchell, R. Gender differences in relationships between urban green space and health in the United Kingdom. Social science & medicine 71, 568–575 (2010).

52 Steptoe, A. & Kivimäki, M. Stress and cardiovascular disease. Nature Reviews Cardiology 9, 360–370 (2012).

53 Pollitt, R. A., Rose, K. M. & Kaufman, J. S. Evaluating the evidence for models of life course socioeconomic factors and cardiovascular outcomes: a systematic review. BMC public health 5, 7 (2005).

54 Phillips, S. P. Defining and measuring gender: a social determinant of health whose time has come. International Journal for Equity in Health 4, 1–4 (2005).

55 Bishop, C. M. Bayesian methods for neural networks. (1995).

56 Dreiseitl, S. & Ohno-Machado, L. Logistic regression and artificial neural network classification models: a methodology review. Journal of biomedical informatics 35, 352–359 (2002).

57 Smith, K. P. & Christakis, N. A. Social networks and health. Annu. Rev. Sociol 34, 405–429 (2008).

58 Suglia, S. F. et al. Why the neighborhood social environment is critical in obesity prevention. Journal of Urban Health 93, 206–212 (2016).

59 Bahr, D. B., Browning, R. C., Wyatt, H. R. & Hill, J. O. Exploiting social networks to mitigate the obesity epidemic. Obesity 17, 723–728 (2009).

60 Pachucki, M. A., Jacques, P. F. & Christakis, N. A. Social network concordance in food choice among spouses, friends, and siblings. American journal of public health 101, 2170–2177 (2011).

61 Årnes, A. P. & Krokstrand, T. T. The incidence and prevalence of Chronic Fatigue Syndrome, Back Pain of unknown origin, Fibromyalgia, and Myalgia in Norwegian women, and their association to physical activity. A prospective cohort study of material from the Norwegian Women and Cancer (NOWAC) study, UiT Norges arktiske universitet, (2014).

62 Ahmad, M. A., Eckert, C. & Teredesai, A. in Proceedings of the 2018 ACM international conference on bioinformatics, computational biology, and health informatics. 559-560.

63 Ni, Y. et al. Towards phenotyping stroke: Leveraging data from a large-scale epidemiological study to detect stroke diagnosis. PloS one 13, e0192586 (2018).

64 Ambale-Venkatesh, B. et al. Cardiovascular event prediction by machine learning: the multi-ethnic study of atherosclerosis. Circulation research 121, 1092–1101 (2017).

65 Sitar-tăut, A., Zdrenghea, D., Pop, D. & Sitar-tăut, D. Using machine learning algorithms in cardiovascular disease risk evaluation. Age 1, 4 (2009).

66 Weng, S. F., Reps, J., Kai, J., Garibaldi, J. M. & Qureshi, N. Can machine-learning improve cardiovascular risk prediction using routine clinical data? PloS one 12, e0174944 (2017).

67 Bazemore, A. W. et al. “Community vital signs”: incorporating geocoded social determinants into electronic records to promote patient and population health. Journal of the American Medical Informatics Association 23, 407–412 (2016).

68 Cantor, M. N. & Thorpe, L. Integrating data on social determinants of health into electronic health records. Health Affairs 37, 585–590 (2018).

